# Suicide prevention curriculum development for health and social care students: Protocol for a scoping review

**DOI:** 10.1101/2023.04.19.23288793

**Authors:** Kerrie Gallagher, Clíodhna O’ Brien, Doireann Ní Dhalaigh, Paul Corcoran, Eve Griffin

**Author notes:** Corresponding author (EG). **Funding** This study was funded by the National Office for Suicide Prevention, Health Service Executive, Ireland. **Competing interests** The authors declare they have no competing interests. **Data availability** For requests to access data gathered from this review, the corresponding author can be contacted.

## Abstract

**Background:** Suicide has become a serious public health concern and international research has shown that the majority of individuals who died by suicide had received healthcare in the year prior to their death. This presents an opportunity for suicide prevention by strategically training healthcare students in suicide prevention knowledge and skills.

**Objective:** The objective of this scoping review is to identify the literature on the development and implementation of suicide prevention curricula for undergraduate and postgraduate students of health and social care degree programmes.

**Inclusion criteria:** Studies will only be considered eligible for inclusion if they describe the development and/or implementation of suicide prevention curricula being taught to health and social care degree students in higher education/university settings. Quantitative, qualitative and mixed method studies published between 2011 and 2023 (inclusive) and in the English language will be considered eligible.

**Methods:** This scoping review will be conducted according to the PRISMA guidelines for scoping reviews (PRISMA-ScR). The developed search strategy will be implemented across seven databases: Australian Educational Index, British Educational Index, ERIC (Education Resources Information Center), PsycINFO, Embase, Scopus and Web of Science. Several grey literature databases will also be consulted. Further potential results will be located by hand-searching the reference lists of included articles. The search strategy will include variations of the terms ‘university students’, ‘suicide prevention’ and ‘education’. The search terms will be limited to titles, abstracts, and keywords in databases that allow it. Two reviewers will complete the screening using the predefined inclusion criteria. A third reviewer will resolve any conflicts during the screening and eligibility appraisal processes. Results will be presented in the form of tabulated results and an accompanying narrative summary, describing key findings and context related to learning outcomes, methodologies employed and implementation of the identified programmes.

## Introduction

It is widely recognised that suicide is a serious public health concern. Healthcare settings provide a unique opportunity to incorporate suicide prevention strategies (1). These strategies can focus on providing mental health support, screening for suicidal risk, early intervention, and creating an environment that encourages help-seeking behaviour (2,3). However, evidence shows that many existing clinical training programmes do not adequately prepare healthcare professionals to engage in suicide prevention (4,5).

A population-based cohort study conducted in Sweden between 2004 and 2015 found that 86% of individuals who died by suicide received healthcare in the year preceding their death (6). It was also estimated that in the month before their death, 53% of those who died by suicide had received some form of healthcare consultation. This is compared with 20% in the general population (6).

Health professionals, such as nurses, pharmacists, primary care providers and social workers, are well-positioned to detect signs and symptoms of suicidal ideation and behaviours and provide those at risk with the appropriate referrals to support (7–10). Training of healthcare staff has been highlighted as an important element of suicide intervention and prevention (11). However, healthcare staff often do not receive appropriate training or lack knowledge and awareness of suicide and self-harm that allows for an appropriate response to suicide risk, primarily due to a lack of resources for healthcare professionals to gain access to appropriate information and training (12–14). This is reflected in the fact that both professional practitioners and students rate their own interpersonal suicide prevention skills as poor (15). This lack of confidence on the part of healthcare professionals in their own competence in risk screening and intervention may serve as a barrier to effective suicide prevention in the healthcare setting (16,17).

A suicide prevention programme targeted at healthcare students provides an ideal opportunity to ensure a universal approach to suicide prevention in the healthcare system, as all graduates will begin their careers with similar knowledge, attitudes, and skills. To the best of the authors’ knowledge, the extent of the evidence regarding a suicide prevention curriculum for health and social care students has not been reported on to date. The aim of this article is to describe protocol for a scoping review to map the literature on suicide prevention within undergraduate and postgraduate health and social care curricula. This question was prompted by the recognised need for a national standardised undergraduate suicide prevention curriculum in the Republic of Ireland. It is intended that this scoping review will help inform the development of a suicide prevention curriculum for undergraduate health and social care students in the Irish higher education setting.

## Methods

The reporting items in this protocol comply with the Preferred Reporting Items for Systematic Reviews and Meta-Analyses extension for Scoping Reviews (PRISMA-ScR) guidelines (18) (Appendix 1 in Supplementary Information). We chose to adopt a scoping review approach for this research as the evidence on suicide prevention curricula is still emerging, particularly for health and social care students. A scoping review is an appropriate method for this research since the main aim of this review is to give an overview of the available evidence relating to the development and implementation of suicide prevention curricula in third level institutions internationally. To identify potential similarities and differences between suicide prevention programmes, we intend to review the available evidence regarding the content and delivery of these programmes and, where applicable, their impact on student learning. We used the work of Arksey and O’Malley to design the scoping review protocol (19) and utilised five primary stages to inform the study design: (i) identifying the research question; (ii) identifying relevant studies; (iii) study selection; (iv) charting data; (v) collating, summarizing, and reporting the results.

### (i) Identifying the research question

The scoping review will seek to address the following question:

“What educational methodologies are employed to implement suicide prevention courses in higher education, both in terms of content and delivery, and, where applicable, their impact on student learning?”.

#### Eligibility criteria

##### Participants and context

Studies that examine suicide prevention curricula being taught to undergraduate/postgraduate health and/or social care degree students in a higher education setting will be considered eligible for inclusion in the review process. There will be no limits set on region of publication, however, due to limited resources, studies published in the English language since 1^st^ January 2011 will be included for examination. Studies will not be included if they report on suicide prevention training implementation outside of or in addition to the university/higher education curriculum.

##### Types of Sources

Quantitative, qualitative and mixed method studies, as well as any studies that describe the development of a suicide prevention module in higher education settings will be considered for inclusion. Any relevant grey literature will also be included. Table 1 details the inclusion and exclusion criteria for the review.

**Table 1:** Inclusion and exclusion criteria

| Table 1: Inclusion and exclusion criteria |  |
| --- | --- |
| Include | Exclude |
| Articles focused on developing and/or implementing suicide prevention programmes as part of the higher education curriculum | Articles not published in the English language |
| Articles reporting on empirical research | Articles reporting suicide prevention training delivered to health or social care students independent of the third level curriculum |
| Articles reporting on qualitative research | Articles reporting research or programmes for other populations (e.g., GP gatekeeping training) |
| Articles employing mixed methods design |  |
| Descriptive articles detailing the content development and/ or implementation of suicide prevention education in higher education |  |
| Articles published since 2011 | Articles published before 2011 |

### (ii) Identifying relevant studies

The search strategy will aim to locate peer-reviewed articles and any relevant grey literature, if available. An initial limited search of Scopus and Web of Science Core Collection was undertaken to identify the potential scope of published articles on the topic. The text words contained in the titles and abstracts of relevant articles, and the index terms used to describe the articles will be used to develop a full search strategy for seven electronic databases: Australian Educational Index, British Educational Index, ERIC, PsycINFO, Embase, Scopus and Web of Science Core Collection (see Table 2). It is anticipated that the search strategy for the review, including all identified keywords will be tailored to each database. Further to this, relevant grey literature databases will be searched to retrieve items that would not otherwise be available through the above databases. There will be several databases included in this project, including Google Scholar, the Suicide Prevention Resource Center (SPRC), Grey Matters, Open Grey, and ResearchGate. The search terms will be limited to titles, abstracts, and keywords in databases that allow it. It is also planned to manually search the reference lists of the included studies to identify any potentially overlooked materials. Should conference abstracts and study protocols satisfy the inclusion criteria, we will endeavour to retrieve the full article reporting on the development and/or implementation of the curriculum. The search strategy will include variations of the terms ‘*university students’*, ‘*suicide prevention’* and ‘*education*’. An example of the search strategy to be used is displayed in Table 2.

**Table 2:** Search strategy for Web of Science Core Collection

| Database | Rationale for searching | Search strategy |
| --- | --- | --- |
| Web of Science Core Collection | Considering that the Web of Science has multidisciplinary contents from a broad range of academic disciplines, it will be searched. | ((“suicide prevention training”) OR (“suicide prevention education”) OR (“suicide prevention teaching”)) AND ((“higher education”) OR (“third level education”) OR (“college” OR “university”))<br>Limit to English Language<br>Limit to studies published between 2011-2023 |

### (iii) Study selection

Following the search, all identified citations will be collated and uploaded into the reference manager Zotero, Version 6.0.20. After this, all citations will be imported into Rayyan, and duplicates removed. Rayyan is a free web-tool designed to assist researchers conducting systematic reviews, scoping reviews, and other projects involving knowledge synthesis (20). Screening will follow two stages: (i) title and abstract screening; (ii) full-text screening. Regarding the grey literature, which often lacks abstracts, texts will be screened through executive summaries, table of contents, or other comparable methods. If these methods of screening are not available, grey literature will be advanced to full-text screening. Two reviewers will independently conduct both screening stages and assess each stage against the inclusion criteria. In the event of any discrepancies, a third reviewer will be consulted. This search and study inclusion process will be described in full in the final scoping review along with a Preferred Reporting Items for Systematic Reviews and Meta-analyses extension for scoping review (PRISMA-ScR flow chart) (18). Using the PRISMA diagram, the review process will be illustrated and stages where studies were eliminated will be recorded.

### (iv) Charting the data

A purpose-designed data extraction spreadsheet will be used to extract the following information from all included studies: Data items will include details of the publication e.g., author, country, year of publication, study setting (e.g., university,); the study population (degree course, stage of the degree, number of students); details of the programme (e.g., session length, number of sessions); learning outcomes; accreditation body, was the training deemed to be ‘essential’ and was attendance ‘mandatory’; level of content delivered (i.e., foundational, basic, advanced, expert, other, unspecified); how the programme was implemented; methodologies used; staff training needs and assessment methods employed). A draft extraction form is provided (see Appendix 2 in Supplementary Information). The draft data extraction tool will be modified and revised as necessary during the process of extracting data from the various evidence sources that have been included in the review. Any modifications made during this stage of the process will be detailed in the scoping review. If disagreements arise between the reviewers, they will be resolved by involving the third reviewer.

### (v) Collating, summarising, and reporting the results

The objective of this scoping review is to map existing evidence regarding the development and implementation of suicide prevention programmes in higher education. Data abstraction will be conducted by two reviewers independently from full-text records included in the review. As a means of ensuring accuracy and consistency, each reviewer’s abstracted data will be compared, and discrepancies will be discussed with the third reviewer. A request for missing or additional information will be made to the authors of selected records, if necessary. To facilitate the mapping of common patterns in the data, thematic analysis methods will be employed. This process will provide a summary of key findings regarding suicide prevention for health and social care students, including programme design and/or implementation. Where applicable changes in student knowledge, attitudes and confidence will also be reported. Based on the eligibility criteria, it is anticipated that some of the identified studies will be descriptive in nature and will not be subject to critical appraisal; where possible, critical appraisal will be conducted for effectiveness studies. A detailed description of the tools and strategies used for the critical appraisal will be included in the supplementary information of the review, as well as a discussion of its findings in the main text. Any relevant abstracted information will be charted in the form of tabular results and an accompanying narrative summary, which will include key findings and contextual information regarding the topics, learning outcomes, and methodologies employed.

## Limitations

The authors recognize that this study will have limitations. As we are constrained by resources, only studies published in the English language will be included. We have also noted a limitation with respect to the search of grey literature; even if grey literature is reviewed systematically, some sources may be overlooked. We will attempt to overcome this by searching as many sources as possible in the time we have allocated to complete the review.

## Discussion

The interaction between patients and their healthcare professional provides an opportunity to screen and assess for patient suicide risk given the evidence that the majority of people who die by suicide have been in contact with a health professional in the year preceding their death (1). The provision of suicide prevention training for undergraduate students of the healthcare professions presents an ideal opportunity to equip healthcare professionals with relevant suicide prevention skills, knowledge and confidence. The purpose of the proposed review is to give an overview of suicide prevention programmes that are taught within undergraduate and postgraduate health and social care curricula. The information gathered will be presented in a published scoping review.

Seven databases have been identified as being most relevant to the present study. Two independent reviewers will review the abstracts and full texts of the search to identify eligible and relevant papers which will contribute to our understanding of suicide prevention in higher education.

As opposed to consulting with stakeholders at the end of the scoping process, our partners will be involved at various stages of the process. The findings will be used by the authors in National Suicide Research Foundation with the Health Service Executive National Office for Suicide Prevention to inform the development of a national suicide prevention curriculum for all health and social care undergraduate students. The findings will provide an evidence-base that can be utilised to develop and/or further enhance suicide prevention curricula in higher education institutions in the Republic of Ireland.

## Data Availability

No datasets were generated or analysed during the current study. All relevant data from this study will be made available upon study completion.

## Authors’ contributions

Conceptualisation: Kerrie Gallagher, Clíodhna O’ Brien, Doireann Ní Dhalaigh, Paul Corcoran, Eve Griffin

Funding Acquisition: This study was commissioned by the Health Service Executive, National Office for Suicide Prevention (NOSP).

Methodology: Kerrie Gallagher, Clíodhna O’ Brien, Doireann Ní Dhalaigh Paul Corcoran, Eve Griffin Project administration: Kerrie Gallagher, Clíodhna O’ Brien, Doireann Ní Dhalaigh

Writing – original draft: Kerrie Gallagher, Clíodhna O’ Brien, Eve Griffin

Writing – review and editing: Kerrie Gallagher, Clíodhna O’ Brien, Doireann Ní Dhalaigh, Paul Corcoran, Eve Griffin

## Acknowledgements

The authors would like to acknowledge the support of the Health Service Executive National Office for Suicide Prevention, in particular Prof. Phillip Dodd, Clinical Advisor and Ms. Ailish O’Neill, National Education and Training Manager.

## Notes

### Competing Interest Statement

The authors have declared no competing interest.

### Funding Statement

1. KG, COB, DND, PC, EG 2. N/A 3. Health Service Executive, National Office for Suicide Prevention 4. https://www.hse.ie/eng/services/list/4/mental-health-services/nosp/ 5. No, the National Office for Suicide Prevention The funders did not and will not have a role in study design, data collection and analysis, decision to publish, or preparation of the manuscript.

